# Genetic ancestry proportion influences risk of adverse events from tuberculosis treatment in Brazil

**DOI:** 10.1101/2024.08.29.24312595

**Authors:** Jacqueline A. Piekos, Gustavo Amorim, Felipe Ridolfi, Marcelo Cordeiro-Santos, Afrânio L. Kritski, Marina C. Figueiredo, Bruno B. Andrade, Adalberto R. Santos, David W. Haas, Timothy R. Sterling, Valeria C. Rolla, Digna R. Velez Edwards

## Abstract

Tuberculosis (TB) treatment is highly effective, but response to therapy can vary by geography, race, and ethnicity. We assessed for differences in TB treatment response in a representative and heterogeneous Brazilian population. We estimated genetic ancestry proportion according to major ancestry groups (African, European, and Amerindian ancestry) in the Regional Prospective Observational Research in Tuberculosis (RePORT)-Brazil cohort. RePORT-Brazil is an observational prospective cohort study of individuals with newly-diagnosed, culture-confirmed, pulmonary TB. TB treatment outcomes that were attributed to TB treatment included Grade 2 or higher adverse drug reaction (ADR), Grade 3 or higher ADR, hepatic ADR, and failure/recurrence. Ancestry proportion was the main predictor in logistic regression for each outcome, with adjustments for candidate confounders. There were 941 pulmonary TB patients included in this study. We observed a decreased risk of Grade 2+ ADR when African ancestry proportion increased by 10% (Odds Ratio [OR] 0.41, 95% Confidence Interval [CI] 0.20 -0.85) and an increased risk for Grade 2+ ADR with increasing European ancestry (OR 2.84, 95% CI 1.47 - 5.48). We then performed the same analysis adding HIV status as an interaction term. The decreased risk for Grade 2+ ADR seen for African ancestry proportion did not hold for persons living with HIV; we observed increased risk for Grade 2+ ADR with increasing African ancestry proportion. There were no associations with Amerindian ancestry or for other treatment outcomes. In this Brazilian TB cohort, toxicity risk was associated with African and European ancestry, divergent, and affected by HIV.

#RePORT-Brazil Consortium members include:

- Aline Benjamin and Flavia M. Sant’Anna

- Instituto Nacional de Infectologia Evandro Chagas, Fiocruz, Rio de Janeiro, Brazil
- Jamile Garcia de Oliveira and João Marine

- Clínica de Saúde Rinaldo Delmare, Rio de Janeiro, Brazil
- Adriana Rezende and Anna Cristina Carvalho

- Secretaria de Saúde de Duque de Caxias, Rio de Janeiro, Brazil
- Michael Rocha and Betânia Nogueira

- Instituto Brasileiro para Investigação da Tuberculose, Fundação José Silveira, Salvador, Brazil
- Alexandra Brito and Renata Spener

- Fundação Medicina Tropical Dr. Heitor Vieira Dourado, Manaus, Brazil
- Megan Turner

- Vanderbilt University Medical Center, Nashville, USA

## INTRODUCTION

Tuberculosis (TB) is a global health problem that remains endemic in places throughout the world. According to the World Health Organization (WHO), Brazil is one of 30 high TB burden countries.[1] In 2022, the incidence of new TB cases in Brazil was 36.3 cases per 100,000 people, while the mortality rate was 2.3 deaths per 100,000 people.[2, 3] Over the last ten years, TB incidence has increased slightly in Brazil, while the mortality rate has remained steady at 2.2 deaths per 100,000 population.[4]

In 2014, WHO presented its “End TB Strategy” with the goal to end the endemic of TB worldwide by 2030. An important pillar of strategy was to treat all TB patients with standard therapy: isoniazid, rifampin, pyrazinamide, and ethambutol for two months, followed by isoniazid and rifampin for four months.[5] Overall, 70% of TB patients treated in Brazil are cured.[6] Over the course of treatment, patients may experience adverse events, including hepatotoxicity. There may also be unsuccessful TB treatment outcomes such as treatment failure or TB recurrence after treatment completion. Co-morbidities such as HIV infection can also affect TB treatment outcome.[7]

Adverse drug reactions (ADR) to TB treatment are common. On average, 5 to 33% of individuals undergoing TB treatment experience adverse events.[8] A previous study performed in Brazil reported adverse events from TB treatment in 11% of patients.[9] Genetics is known to play a role in the drug metabolism of anti-TB treatment—specifically mutations in the *NAT2* gene, which can affect isoniazid metabolism and toxicity.[10] Individuals with two slow acetylator alleles of the *N-Acetyltransferase2* (*NAT2*) gene have been found to be at higher risk of ADR from anti-TB treatment.[10] *NAT2* genotypes are estimated to account for 80% of the pharmacokinetic variability of isoniazid. [11–13] In 2021, Verma et al. proposed a prototype assay for *NAT2* genotype determination with the goal of using it for dose-adjusted isoniazid treatment.[14] Other genetic factors that influence risk for ADR from anti-TB treatment are more poorly understood. Research is ongoing to find biomarkers besides *NAT2* that affect anti-TB treatment outcomes. Using transcriptomics, researchers haves looked for biomarkers that effect TB treatment response [15] and longevity of treatment. [16]

Our present study evaluated genetic ancestry differences in TB drug response— specifically toxicity and effectiveness outcomes—within a Brazilian TB cohort. The population of Brazil has high levels of genetic admixture primarily across European, African, and indigenous ancestry groups, as well as high incidence and prevalence of TB. [17,18] In this study we characterized the ancestry proportion of study participants, and tested for associations between genetic ancestry and TB treatment outcomes.

## MATERIALS AND METHODS

### Cohort Description

The study population was from Regional Prospective Observational Research in Tuberculosis (RePORT)-Brazil, an observational prospective cohort study of individuals with newly-diagnosed, culture-confirmed, pulmonary tuberculosis and their close contacts.[19] Participant enrollment occurred between June 2015 and June 2019 at five sites across three regions of Brazil. Three sites were in Rio de Janeiro (Instituto Nacional de Infectologia Evandro Chagas, Clínica de Saúde Rinaldo Delmare, Secretaria de Saúde de Duque de Caxias), one in Salvador (Instituto Brasileiro para Investigação da Tuberculose), and one in Manaus (Fundação Medicina Tropical Dr. Heitor Vieira Dourado). Characteristics of RePORT-Brazil have been described elsewhere; the study population is representative of all TB cases reported in Brazil.[19]

RePORT-Brazil participants were included in this study if they provided samples for genetic testing and had culture-confirmed, drug-susceptible, pulmonary tuberculosis and received standard anti-TB therapy (isoniazid, rifampin, pyrazinamide, and ethambutol for two months, followed by isoniazid and rifampin for four months). Participants were followed for 24 months to assess TB treatment response and for TB recurrence. RePORT-Brazil excluded individuals who had previously received anti-TB therapy for ≥7 days, received >7 days of fluoroquinolone therapy within 30 days prior to enrollment, were pregnant or breastfeeding, or planned to leave the region during follow-up.

All RePORT-Brazil study participants provided informed consent. The study was approved by the Institutional Review Boards at all study sites and at Vanderbilt University Medical Center.

### Data Collection

Clinical, demographic, and outcome (see below) data, as well as blood, urine, and plasma specimens, were collected longitudinally at four in-person study visits per RePORT-Brazil protocol. The following information was collected from participants at baseline: sex, age, body mass index (BMI), HbA1c, illicit drug use, alcohol use, and tobacco use. All participants were tested for HIV at baseline unless already known to have HIV. The majority of participants were prescribed directly observed therapy (DOT) at treatment initiation.

Seven toxicity outcomes encompassing TB treatment response and recurrence were evaluated in this study. For the outcomes grade 2 or higher adverse drug reaction (ADR), Grade 3 or higher ADR, and hepatic ADR, we tested all cases that had the outcome as well as a nested secondary set of cases that had the outcome attributed to TB treatment. When testing the outcomes attributed to TB treatment, cases who experienced the outcome but it was not attributed to TB treatment, were censored from analysis. Categories of physician-assigned attribution of causality were “possibly”, “likely”, or “definitely” related to TB treatment. The last outcome was treatment failure and/or recurrence. Treatment failure was defined as remaining sputum culture-positive or smear-positive at month 5 or later during treatment. Recurrence cases were defined as culture-confirmed tuberculosis or symptoms consistent with tuberculosis after the participant was considered cured or completed treatment. For case-control analyses, cases were individuals who were recorded as experiencing the outcome and controls were individuals who did not.

### Genotyping

DNA was extracted from whole blood. For ancestry informative markers, genotyping was performed at Laboratório de Biologia Molecular Aplicada a Micobactérias in Fiocruz, using xMAP multiplexing technology (Luminex Corporation). There were 46 ancestry-informative markers (AIMs) selected for genotyping based on a previous analysis of a large and diverse Brazilian cohort.[17] *NAT2* genotyping was performed by VANTAGE (Vanderbilt Technology for Advanced Genomics) using MassARRAY® iPLEX Gold (Agena Bioscience™, California, USA) and Taqman (ThermoFisher Scientific, Massachusetts, USA). Assay design is available upon request. We used *NAT2*[20, 21] gene variants (rs1801280, rs1799930, rs1799931, and rs1801279) to determine drug metabolism status, with intermediate (heterozygous at a single locus), slow (homozygous at any locus or heterozygous at 2 or more loci), and rapid (no variant allele) variants and used this variable as a covariate in analyses.

### Statistical Analysis

ADMIXTURE[22] was used to determine ancestry proportions of each participant based upon the selected AIMs.[23] Using all references from the superpopulations of Europe, America, and Africa (n = 20) from the Human Genome Diversity Project (HGDP),[24] we projected the 46 AIMs onto the reference populations and had unsupervised ADMIXTURE resolve the reference populations into K = 3 groups. ADMIXTURE calculated the allele frequencies of the AIMs for the reference populations and excluded reference populations that either could not be resolved with K = 3 means or were determined to be too heterogenous with ancestry proportions below 60%. Fifteen of the reference populations were resolved into three superpopulations: European (EUR), African (AFR), and Amerindian (AME). The European superpopulation included Adygei in Caucasus, Russia, Basque in France, Bergamo Italian from Italy, Sardinian from Sardinia, Italy, Russians from Russia, and the Orcadian population from Orkney. The African superpopulation included Bantu from Kenya and South Africa, Biaka in Central African Republic, Mandenka in Senegal, Mbuti in Democratic Republic of Congo, and Yoruba from Nigeria. The Amerindian superpopulation included Colombian in Colombia, Karitiani from Brazil, and Maya from Mexico. Using unsupervised ADMIXTURE at several K means, ancestry proportions were determined for the value of K and then projected onto the cohort. K = 3 was the best performing model reconstituting the three superpopulations that were identified in the reference. Ancestry proportions of the three superpopulations were then calculated for all samples.

Logistic regression models were used to evaluate the effect ancestry proportion had on selected outcomes using the “rms” R package in R version 4.0.2.[25] Each of the three superpopulations were used as the main predictor for each outcome, adjusted for age, sex, BMI, study site, HIV status, baseline HbA1c, directly observed therapy (DOT), *NAT2* genotype, and usage of illicit drugs, alcohol, and tobacco. Covariate measurements were taken at baseline enrollment. Outcomes were modeled using ancestry and covariates, ancestry only, and covariates only. The reference group for region of enrollment was Rio de Janeiro and it included all three enrollment sites for that region. For *NAT2,* the rapid genotype was used as the reference for intermediate and slow metabolizers. Samples that did not pass genotyping quality control (QC) or had missing covariate or outcome information were excluded from the analysis.

## RESULTS

### Population Characteristics

Clinical and demographic characteristics of the cohort and case control counts for each outcome evaluated stratified on HIV status is provided in Table 1A-B. The ancestry proportions of all individuals in the cohort are summarized in Figure 1. There were 930 participants who were successfully genotyped and contained all covariates used in the analysis. The most frequent outcome was Grade 2+ ADR (15%), followed by Grade 2+ ADR attributed to TB treatment (11%). (Table 1A) An HIV negative individual had the average ancestry proportions of 42% European, 34% African, and 24% Amerindian while HIV positive individuals were 42% European, 23% African, and 35% Amerindian. (Table 1B) Characteristics according to HIV status including study site, sex, DOT, *NAT2* genotype, and smoking, alcohol, and illicit drug consumption are given in Table 1B. The Manaus site contributed the most HIV-positive participants. Within each HIV stratum, there was a difference in the proportions of *NAT2* genotypes. Among HIV-negative participants, 40% were slow metabolizers and 8% were rapid metabolizers; among HIV-positive, 9% were slow metabolizers and 42% rapid metabolizers (Table 1B).

**Figure 1.**
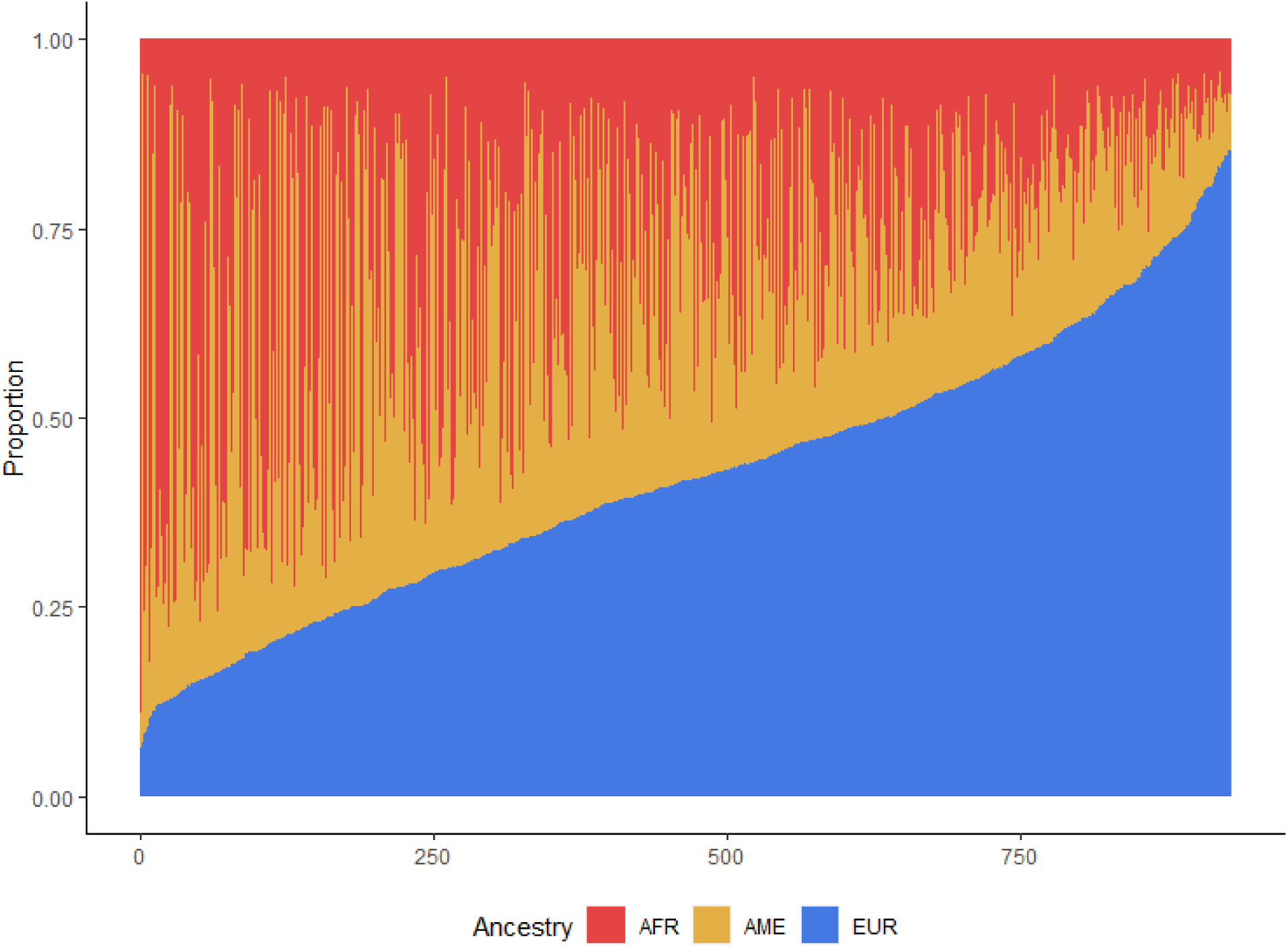
Ancestry proportions of the 930 genotyped study participants from the Brazil RePORT cohort. Subjects are represented on the x-axis and are ordered from lowest to highest proportion of European ancestry. The average person in the cohort had ancestry proportions of 42% European, 32% African, and 26% Amerindian.

**Table 1.** Population characteristics and outcomes among participants in the RePORT Brazil cohort stratified by HIV status.

A. Case control counts for TB treatment outcomes stratified on HIV status
| Outcome | HIV Negative |  | HIV Positive |  |
| --- | --- | --- | --- | --- |
|  | Control* | Case | Control | Case |
| Grade 2+ ADR related to TB treatment | 664 | 62 | 126 | 36 |
| Grade 3+ ADR related to TB treatment | 664 | 18 | 126 | 16 |
| Hepatic ADR related to TB treatment | 730 | 17 | 164 | 16 |
| TB Treatment Failure/Recurrence | 722 | 27 | 170 | 11 |

HIV: human immunodeficiency virus; TB: Tuberculosis; \*Controls excluded individuals who were cases for a specific treatment outcome

B. Demographic characteristics and genetic ancestry estimates
| Variables | HIV Negative<br>(N=749) | HIV Positive<br>(N=181) |
| --- | --- | --- |
| *Categorical Variables* |  |  |
| Site, N (%) |  |  |
| Rio de Janeiro | 379 (51) | 44 (24) |
| Salvador | 229 (31) | 2 (1) |
| Manaus | 141 (19) | 135 (75) |
| Sex, N (%) |  |  |
| Males | 480 (64) | 139 (77) |
| Females | 269 (36) | 42 (23) |
| DOT, N (%) |  |  |
| Not Prescribed | 234 (31) | 44 (24) |
| Prescribed | 515 (69) | 137 (76) |
| NAT2 Genotype, N (%) |  |  |
| Rapid | 60 (8) | 76 (42) |
| Intermediate | 386 (52) | 88 (49) |
| Slow | 303 (40) | 17 (9) |
| Tobacco Use, N (%) |  |  |
| Never (ref) | 373 (50) | 71 (39) |
| Previous user | 194 (26) | 80 (44) |
| Current User | 182 (24) | 30 (17) |
| Alcohol Use, N (%) |  |  |
| Never (ref) | 136 (18) | 12 (7) |
| Previous user | 248 (33) | 108 (60) |
| Current User | 365 (49) | 61 (36) |
| Illicit Drug Use, N (%) |  |  |
| Never (ref) | 530 (71) | 84 (46) |
| Previous user | 128 (17) | 73 (40) |
| Current User | 91 (12) | 24 (13) |
| *Continuous Variables* |  |  |
| BMI, Mean kg/m<sup>2</sup> (SD) | 20.60 (3.56) | 20.61 (1.35) |
| Age, Mean yrs (SD) | 38.36 (15.00) | 36.02 (10.87) |
| HbA1c, Mean (SD) | 6.55 (2.17) | 5.92 (1.15) |
| Genetic Ancestry Proportion, Mean (SD) |  |  |
| African | 0.34 (0.20) | 0.23 (0.17) |
| European | 0.42 (0.18) | 0.41 (0.18) |
| Amerindian | 0.24 (0.15) | 0.35 (0.20) |

HIV: human immunodeficiency virus; ref: reference; DOT: directly observed therapy; SD: standard deviation

### Ancestry Associations

The outcome grade 2+ ADR attributed to TB treatment was significantly associated with African (decreased risk) and European (increased risk) ancestry proportion in adjusted and unadjusted models, while grade 3+ ADR attributed to TB treatment was significantly associated with Amerindian ancestry proportion in the unadjusted analysis, but not the adjusted model. (Table 2) All other outcomes evaluated did not have significant association with ancestry proportion. Increasing African ancestry proportion was associated with decreased risk for grade 2+ ADR attributed to TB treatment while increasing European ancestry proportion was associated with increased risk for grade 2+ ADR attributed to TB treatment. (Figure 2A) For a 10% increase in African ancestry proportion, the OR of having a grade 2+ ADR attributed to TB treatment was 0.41 (95% CI: 0.20 – 0.85) and for a 10% increase in European ancestry, the OR was 2.84 (95% CI: 1.47 – 5.48). (Figure 2B)

**Figure 2.**
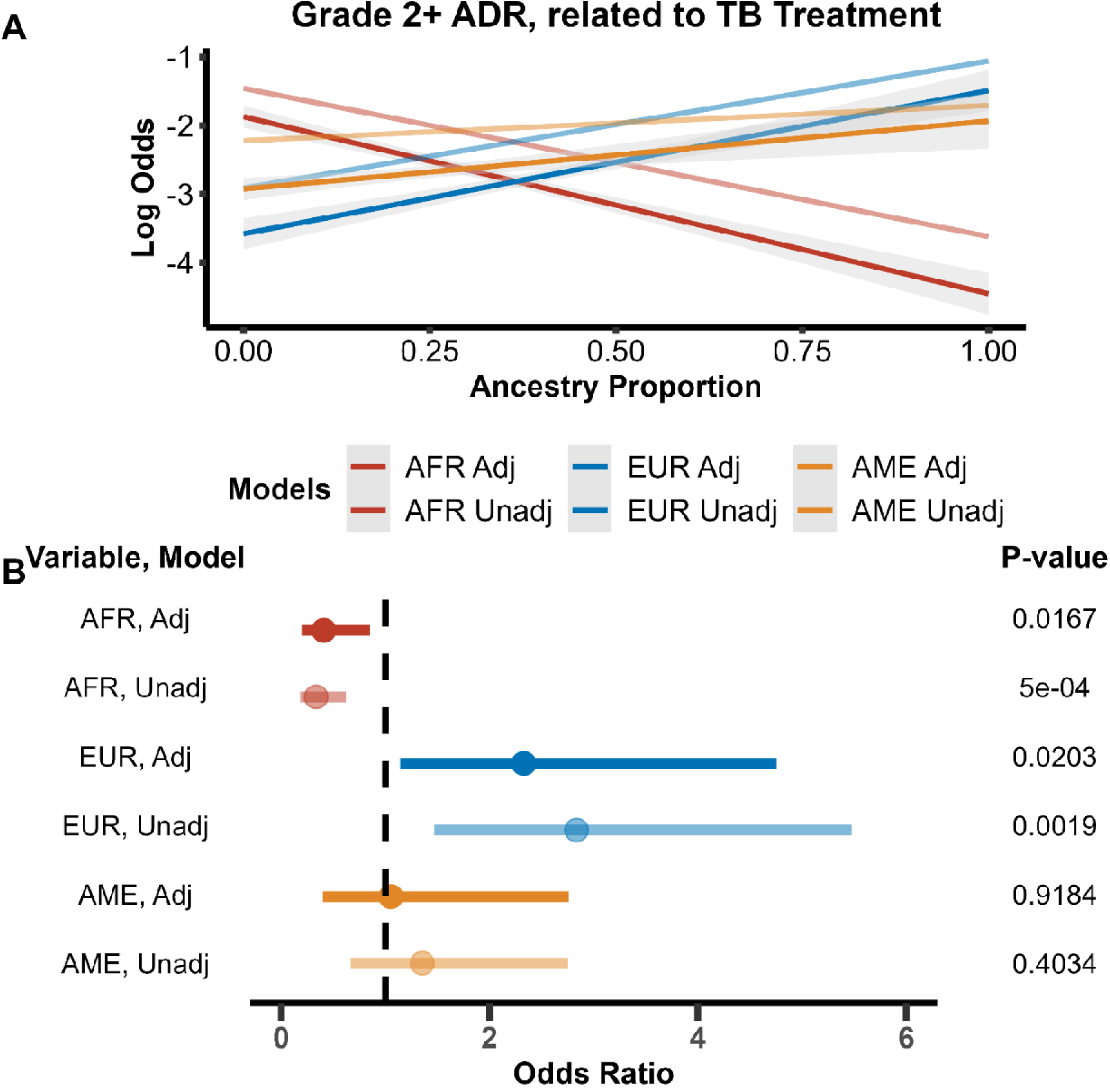
Logistic regression model results of outcome grade 2+ adverse drug reaction (ADR) related to tuberculosis (TB) using ancestry proportion as the main predictor. Unadjusted models are in darker colors, while the lighter colors are the models adjusted for BMI, age, HbA1c, sex, HIV status, DOT status, recruitment site, and smoking, alcohol, and drug usage. A) Unadjusted and adjusted grade 2+ ADR related to TB modeled as log odds of having outcome as a function of ancestry proportion. African and European ancestry were found to be significant predictors for having a grade in the adjusted and unadjusted models and having a general adverse event in the unadjusted models. Increasing African ancestry genetic proportion decreases the risk of having an adverse drug event, related to TB or not, while increasing European ancestry genetic proportion increases the risk. B). Odds ratios of ancestry variables and interaction terms of unadjusted and adjusted models plotted with their standard error. Significance level for variables is listed on the right.

**Table 2.**
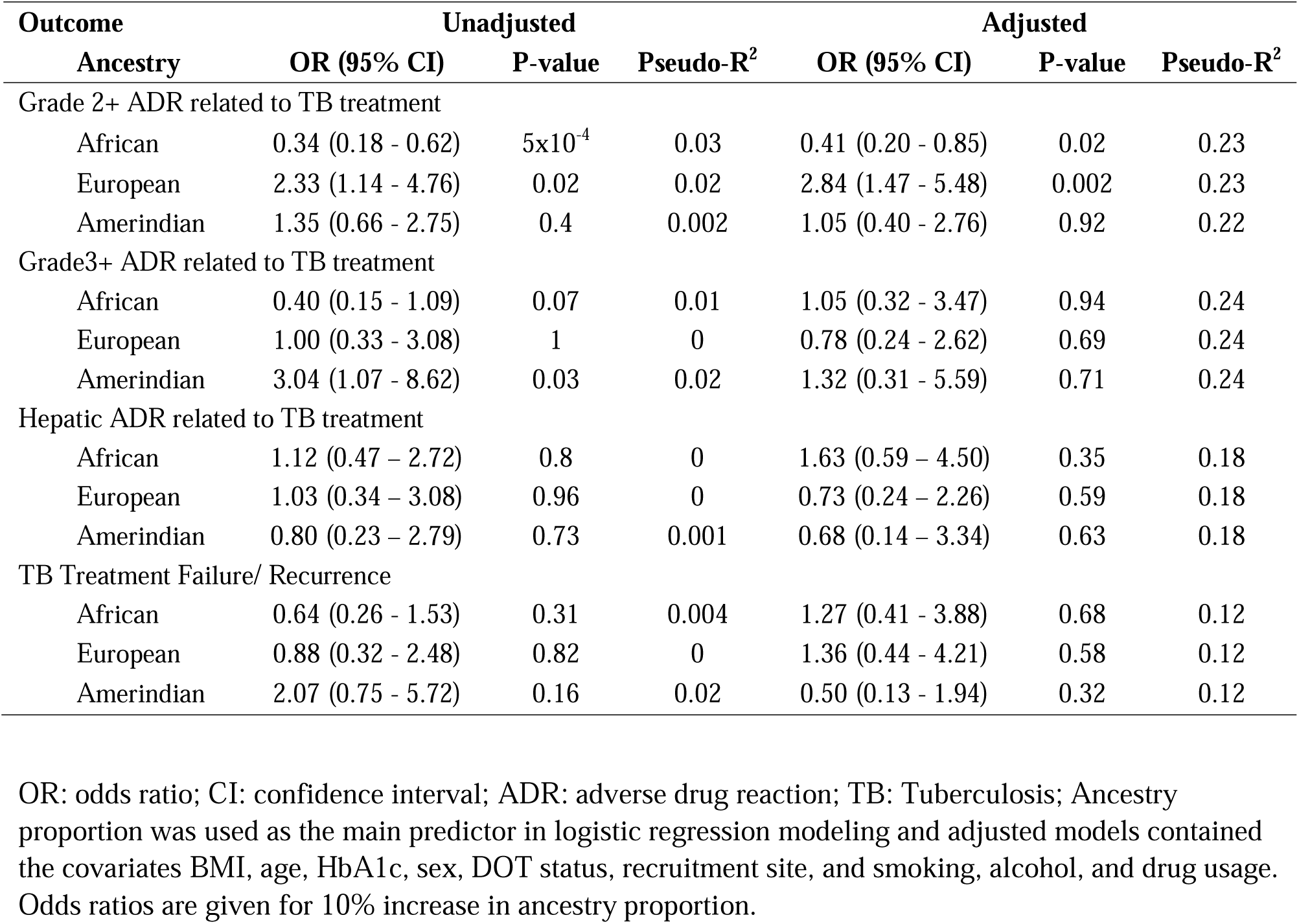
Odds ratio of treatment outcomes for each 10% increase in ancestry proportion and significance level in unadjusted and adjusted logistic regression models.

**Table 3.** Grade 2+ adverse drug reaction related to tuberculosis treatment modeled using logistic regression with an interaction between ancestry proportions and HIV status.

| Variable<br>Interaction Term | Unadjusted |  |  | Adjusted |  |  |
| --- | --- | --- | --- | --- | --- | --- |
|  | OR (95% CI) | P-value | Pseudo-R<sup>2</sup> | OR (95% CI) | P-value | Pseudo-R<sup>2</sup> |
| African | 0.26 (0.13 - 0.52) | 1x10<sup>-4</sup> | 0.10 | 0.36 (0.17 - 0.76) | 0.008 | 0.26 |
| African*HIV | 8.26 (2.64 - 25.79) | 3x10<sup>-4</sup> |  | 4.18 (1.22 - 14.37) | 0.02 |  |
| European | 3.14 (1.55 - 6.38) | 0.002 | 0.08 | 2.17 (1.02 - 4.64) | 0.05 | 0.26 |
| European*HIV | 0.26 (0.08 - 0.89) | 0.03 |  | 0.41 (0.11 - 1.52) | 0.18 |  |
| Amerindian | 1.79 (0.80 - 4.02) | 0.16 | 0.07 | 1.57 (0.57 - 4.28) | 0.38 | 0.25 |
| Amerindian*HIV | 0.33 (0.10 - 1.14) | 0.09 |  | 0.49 (0.13 - 1.87) | 0.29 |  |

OR: odds ratio; CI: confidence interval; ADR: adverse drug reaction; TB: Tuberculosis; Unadjusted models contain ancestry and interaction term. Ancestry proportion was used as the main predictor in logistic regression modeling and adjusted models contained the covariates BMI, age, HbA1c, sex, DOT status, recruitment site, and smoking, alcohol, and drug usage. Odds ratios are given for 10% increase in ancestry proportion.

### Interaction with HIV

We then tested for and modeled an interaction between ancestry proportion and HIV status. When stratified on HIV status we observed opposing associations for HIV negative and HIV positive strata in all three ancestry groups. (Supplemental Table 1A and B). In the HIV negative stratum, increasing African ancestry was associated with decreased risk for grade 2+ ADR while increasing European and Amerindian ancestry were associated with increased risk. (Supplemental Figure 1A) In HIV positive stratum, the opposite association trends were observed. (Supplemental Figure 1B) When ancestry proportion was modeled with an HIV interaction term, increasing African ancestry proportion was associated with a significantly decreased risk for grade 2+ ADR attributed to TB treatment in adjusted and unadjusted models, while increasing European ancestry increased risk for grade 2+ ADR attributed to TB treatment. (Figure 3A) The interaction term for African ancestry and HIV had an odds ratio of 8.26 (95% CI: 2.64 – 25.79, p = 3.0x10^-4^) in the unadjusted model and 4.18 (95% CI: 1.22 – 14.37, p = 0.02) in the adjusted model. The interaction term for European ancestry was 0.26 (95% CI: 0.08 – 0.89, p = 0.03) in the unadjusted model and 0.41 (95% CI: 0.11 – 1.52, p = 0.18) in the adjusted model. (Table 4, Figure 3B) Amerindian ancestry proportion and its interaction with HIV were not significant in unadjusted or adjusted models.

**Figure 3.**
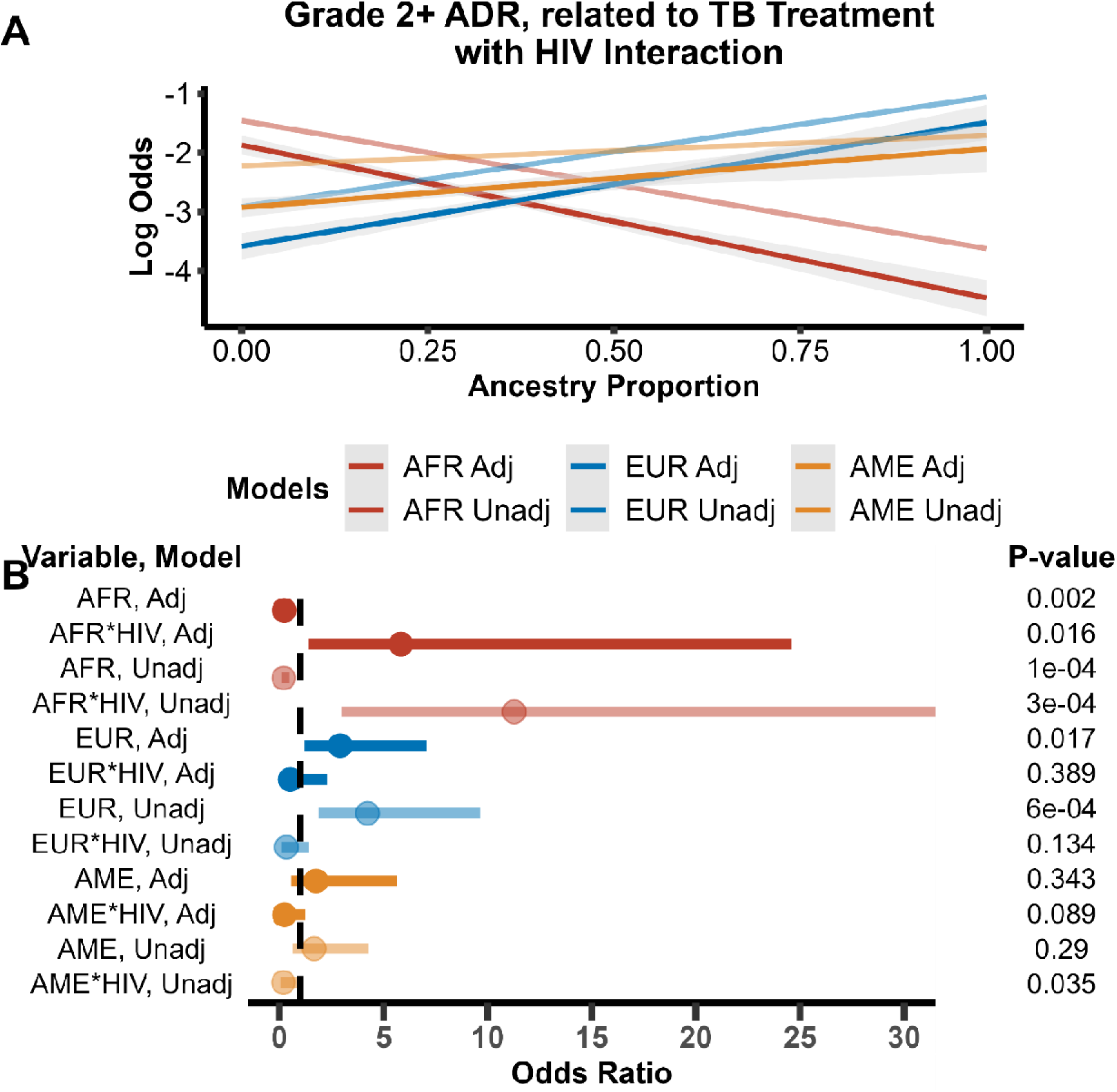
Logistic regression models for grade 2+ adverse drug reaction (ADR) related to TB stratified on HIV status. Adjusted models contain the covariates age, sex, BMI, treatment site, HgA1c, NAT2 genotype, directly observed therapy, and alcohol, smoking and illicit drug use status. A) Unadjusted and adjusted logistic regression models of HIV negative individuals. Increasing African ancestry proportion lowers odds of a grade 2+ ADR related to TB while increasing European and Amerindian ancestry proportion increase odds. B) Unadjusted and adjusted logistic regression models of HIV positive individuals. In this strata, increasing African and Amerindian ancestry proportion lowers odds of a grade 2+ ADR related to TB while increasing European ancestry proportion increase odds.

## DISCUSSION

Here we observed how genetic ancestry influenced the outcomes of TB treatment in a cohort that is reflective of the TB population in Brazil. The average genetic ancestry observed in the cohort was split almost evenly amongst European, African, and Amerindian ancestry. We observed that individuals from the Manaus site had a slightly higher proportion of Amerindian ancestry on average than individuals from Rio de Janeiro and Salvador. This aligns with previous studies in other South American populations that have found individuals in urban areas to have higher proportions of European and African genetic ancestry, and individuals living more inland have a higher proportion of Amerindian genetic ancestry.[26–28]. The Manaus site is further inland in Brazil, where there has been less admixture between the indigenous populations who originally inhabited the area and incoming settlers.

In the RePORT population, we found African genetic ancestry to be protective against grade 2+ ADR attributed to TB treatments while European ancestry was associated with increased risk. The differences in risk we observed cannot all be attributed to the *NAT2* genotypes. This pattern further applies to all grade 2+ ADR. A previous study of ADR in the TB registry in Brazil found self-determined Black and White race to be protective against ADR when compared to Indigenous.[29] This study found African genetic ancestry to decrease risk for a grade 2+ ADR from TB treatments, exemplifying the differences between race and ancestry. The results of these two studies together suggest the racial disparities observed in outcomes may be due to exogenous environmental variables that disproportionately affect self-reported Black race individuals.

TB is known to be a common comorbidity to individuals living with HIV. We found HIV status to be a risk modifier for African genetic ancestry. Despite an overall trend of African genetic ancestry decreasing risk for a grade 2+ ADR due to TB treatment, for individuals living with HIV, increasing African genetic ancestry proportion was associated with an increased risk for a grade 2+ ADR due to TB treatments. In other words, the protective effect observed for African genetic ancestry was negated in individuals living with HIV. Individuals living with HIV were at higher risk for a grade 2+ ADR from TB treatment, regardless of genetic ancestry. There were several limitations of our study.

To our knowledge this is the first study to evaluate the relationship between genetic ancestry and TB treatment outcomes. There has been a previous study that found self-determined race by skin color (Black or Brown compared to White), found that Black and Brown race had increased risk for adverse events from TB.[30] By looking at genetic ancestry, we were able to tease apart contributions of genetics versus environments towards the outcomes studied. We found African genetic ancestry to be protective against adverse TB treatment toxicity outcomes, especially for individuals living without HIV. These data support that disparities in TB treatment outcomes are in part due to differences in genetic ancestry across individuals.

## Data Availability

All data produced in the present study are available upon reasonable request to the authors

## CONTRIBUTORS

Conception of project by JAP, DRVE, TRS, DWH, ARS, BBA, VR. Funding secured by TRS, VCR, DWH, DRVE. Acquisition of data done by VCR, MCS, ALK, BBA, ARS, Analysis of data performed by JAP, DRVE, GA. Writing done by JAP with editing and input from DRVE, TRS, DWH, GA, MCS, ALK, MCF, BBA, ARS.

## ACKNOWLEDGEMENTS

We would like to thank the study participants of RePORT Brazil for their participation. We would like to thank Jacob Keaton for previous work on estimating ancestry proportions for this study. This work was supported by the Departamento de Ciência e Tecnologia, Secretaria de Ciência e Tecnologia, Ministério da Saúde, Brazil (25029.000507/2013-07); National Institute of Allergy and Infectious Diseases (NIAID) (U01 AI069923, R01 AI120790, U01 AI172064, R01 AI077505, P30 AI110527) and the National Center for Advancing Translational Science (UL1 TR000445) at the National Institutes of Health. JAP was funded by T32-GM080178.

## DATA SHARING STATEMENT

Summary data available by request.

## SUPPLEMENTAL

**Supplemental Table 1.** Grade 2+ adverse drug reaction (ADR) related to tuberculosis treatment modeled stratified by HIV status.

A. HIV Negative
| Ancestry | Unadjusted |  |  | Adjusted |  |  |
| --- | --- | --- | --- | --- | --- | --- |
|  | OR (95% CI) | P-value | Pseudo-R<sup>2</sup> | OR (95% CI) | P-value | Pseudo-R<sup>2</sup> |
| African | 0.21 (0.09 - 0.47) | 1x10<sup>-4</sup> | 0.05 | 0.25 (0.10 - 0.64) | 0.004 | 0.24 |
| European | 4.27 (1.87 - 9.76) | 6x10<sup>-4</sup> | 0.04 | 3.06 (1.23 - 7.80) | 0.02 | 0.23 |
| Amerindian | 1.77 (0.62 - 5.09) | 0.29 | 0.003 | 1.66 (0.41 - 6.78) | 0.48 | 0.21 |

B. HIV Positive
| Ancestry | Unadjusted |  |  | Adjusted |  |  |
| --- | --- | --- | --- | --- | --- | --- |
|  | OR (95% CI) | P-value | Pseudo-R<sup>2</sup> | OR (95% CI) | P-value | Pseudo-R<sup>2</sup> |
| African | 2.74 (0.78 - 9.58) | 0.12 | 0.02 | 1.01 (0.20 - 5.01) | 1 | 0.21 |
| European | 1.42 (0.45 - 4.47) | 0.54 | 0.003 | 1.38 (0.41 - 4.63) | 0.60 | 0.21 |
| Amerindian | 0.38 (0.13 - 1.06) | 0.06 | 0.04 | 0.65 (0.17 - 2.49) | 0.53 | 0.21 |

OR: odds ratio; CI: confidence interval; ADR: adverse drug reaction; TB: Tuberculosis; Ancestry proportion was used as the main predictor in logistic regression modeling and adjusted models contained the covariates BMI, age, HbA1c, sex, DOT status, recruitment site, and smoking, alcohol, and drug usage. Odds ratios are given for 10% increase in ancestry proportion.

**Supplemental Figure 1.**
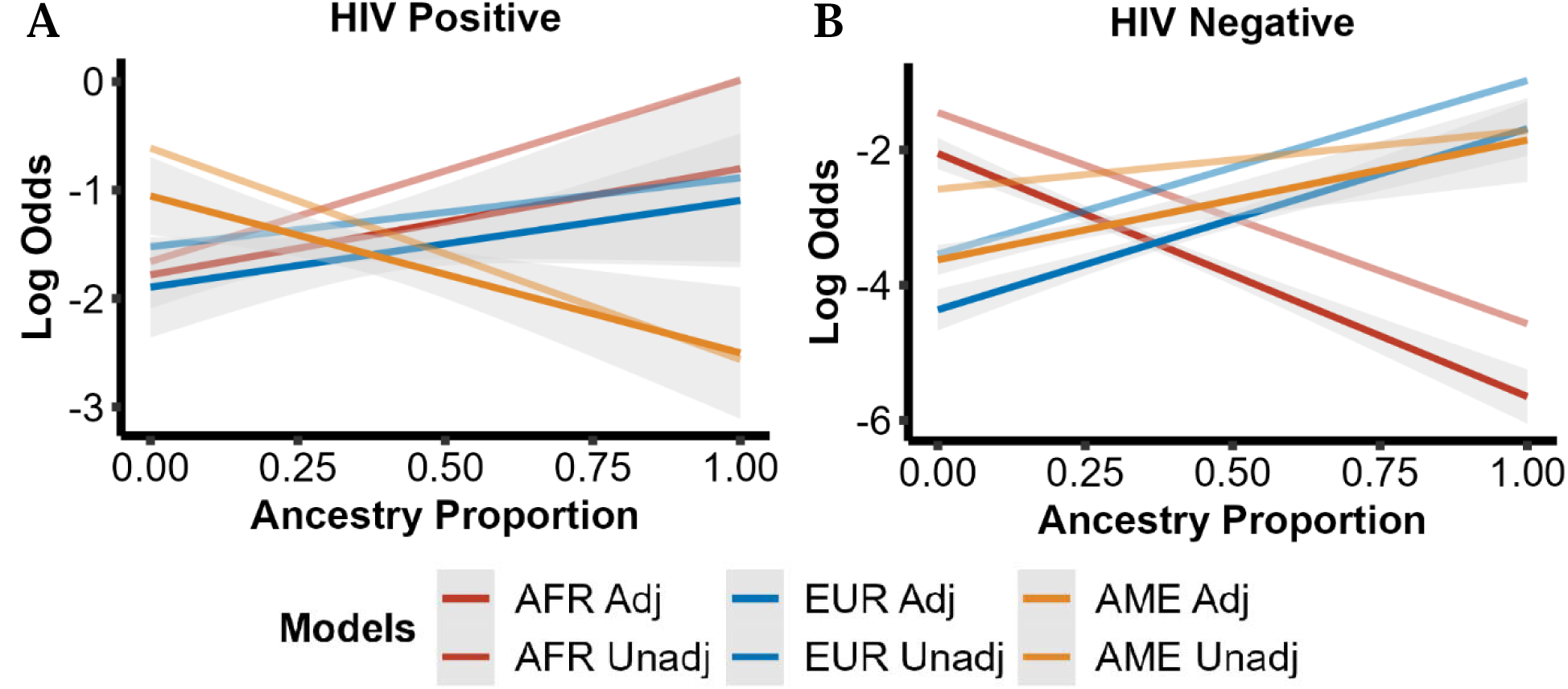
Logistic regression models for grade 2+ adverse drug reaction (ADR) related to TB stratified on HIV status. Adjusted models contain the covariates age, sex, BMI, treatment site, HgA1c, NAT2 genotype, directly observed therapy, and alcohol, smoking and illicit drug use status. A) Unadjusted and adjusted logistic regression models of HIV positive individuals. Increasing African ancestry proportion increases odds of a grade 2+ ADR related to TB while increasing European and Amerindian ancestry proportion decrease odds. B) Unadjusted and adjusted logistic regression models of HIV negative individuals. In these strata, increasing African and Amerindian ancestry proportion increases odds of a grade 2+ ADR related to TB while decreasing European ancestry proportion increase odds.

